# COVID-19 Sniffer Dog experimental training: which protocol and which implications for reliable identification?

**DOI:** 10.1101/2021.06.02.21257981

**Authors:** Silvia Angeletti, Francesco Travaglino, Silvia Spoto, Maria Chiara Pascarella, Giorgia Mansi, Marina De Cesaris, Silvia Sartea, Marta Giovanetti, Marta Fogolari, Davide Plescia, Massimiliano Macera, Raffaele Antonelli Incalzi, Massimo Ciccozzi

**Author notes:** Corresponding Author Unit of Clinical Laboratory Science, University Campus Bio-Medico of Rome, Via Alvaro del Portillo 200, 00128 Rome, Italy.

## Abstract

The introduction of trained sniffer dogs for COVID-19 disease detection could be an opportunity, as previously described for other diseases. Dogs could be trained to detect volatile organic compounds (VOCs), the whiff of COVDI-19 disease. Dogs involved in the study were three one male and two females from different breeds, Black German Shepherd, German Shepherd and Dutch Shepherd. The training was performed using sweat samples from COVID-19 positive apteints and from covid-19 free patients admitted at the University Hospital Campus Bio-medico of Rome. Gauze with sweat were collected in glass jar with metal top and put in metal boxes used for dog training. The dog training protocol was performed in two phase: the olfactory conditioning and the olfactory discrimintaion research. The training palnning was focused on the switch moment for the sniffer dog, the moment when the dog was able to identify VOCs specific for COVID-19 disease. At this time the dog was able to identify VOCs specific for COVID-19 disease with significant reliability, in terms of number of correct versus uncorrect (p<0.0001) reporting. In conclusion, this protocol could provide a useful tool for sniffer dogs training and their introduction in mass screening context, cheaper and faster than a conventional testing method.

## INTRODUCTION

The dog olfactory system with the olfactory mucosa of the nasal cavity have been largely studied for its unique charactersitic consisting in the presence of basal cells providing the regular regeneration of olfactory sensory neurons (OSNs)^1^ and for its abundance of olfactory recepetor reaching about 200 million.^2,18^ The dogs olfactory properties have been largely employed for the reaserach and detection of explosive substances or dead bodies, as Dog support units (DSUs) in police, army and civil protection divisions, in harbours and airport, in private security agency.

The use of sniffer dogs in medical settings can be dated back to 1989 and since then many other applications have been described, such as breast and lung cancers with percentage of detection rate ranging from 88% to 99%^4^ malaria disease with DR of about 82%^5^ and viral or bacterial infections with DR from 77% to 92.6%^6-8^.

Several volatile organic compounds (VOCs) accounts for the odour released by expiration phase of breathing, skin emanation, urine and breath vapors, saliva and pathological conditions. These odours depends on biochemical modification occurring in the body with the consequent release of these specific compounds that are volatile^9^. The metabolic changes occurring in the body in presence of specific conditions such as inflammation, infections or neoplastic disease can be recognised by the dogs that are provided by a powerful olfactory apparatus if adequately trained to the detection of the VOCs.^10^

The same approach could be used for COVID-19 detection, as described in previous studies. ^11-14^ The VOCs could be useful in clinical diagnosis od different disease inclusding bacterial and viral infections as SARS-CoV-2 causing COVID-19interstizial bilateral pneumonia^11-14^. In a recent study, dogs professionally trained were evalutaed for glucose levels detection in patients with diabetes. This study suggested that dogs after adequate training have the ability to detect hypo and hyperglycemic conditions. ^15^

VOCS detection trained dogs could provide early detection of SARSCov-2 infected patients at low cost. The trained dog has the ability to screen more than 200 individuals per hour, enough to allow mass screening at airports, stadium, or in case of crowded events where the virus transmission control by asymptyomatic individuals is fundamental. This is in agreement with World Health Organization (WHO) recommendation about mass screening and its application also in low-income countries where the use of sophisticated and expensive screening tools could be limiting.

The study aims to evaluate the sniffer dogs ability to discriminate VOCs emanated by skin in course of COVID-19 disease, demonstrating that this disease is characterized by a specific odour and that dogs are able to indentify it efficiently and fastly.

## 2 MATERIALS and METHODS

### 2.1 Experimental design

The training planning was developed involving dogs from different breeds. Dogs involved in the study were from different working dog breeds, since their features are useful to standardize the characteristics, the management and the training coherence, to the advantage of more homogeneity in results recording. The intention was to concentrate the experiment on high quality rather than on great number of dogs. Infact, dogs were selected for their specific talents suitable to this kind of experimental design such as temperament, docility and resistance.

Covid-19 conditioned dogs, once involved in the study, will be recoverted to other activities of safety and security, if necessary or at the end of the pandemic, to guarantee the service continuity and mental and physical dogs health in the future. The study was approved by the Local Ethic Committee of University campus Bio-Medico of Rome (PAR 17.21 OSS)

The dogs training plan was divided in two steps: first step of “specific conditioning” to Covid-19 VOCs, consisting in the association of the odour reserch and consequent reporting. This critical and fundamental step is developed using several sweat samples from patients admitted to the Covid Center of the University Hospital Campus Bio-Medico of Rome, for Covid-19 disease. The second step of “olfactory discrimination research” consisited in the discrimination between the COVID-19 odour of interest and everything else that has to be extinguished. At this aim, the discrimination was performed between a box containing underarm sweat collected on gauze from Covid-19 positive patients, a box containing underarm sweat collected on gauze from Covid-19 negative patients, a box containing blank gauze and an empty box. The different boxes were randomly positioned in a line-up from a minimum of four possibilities upwards.

The training involved the repetion of different experimental sessions in line-up to fix more an more the VOCs in the olfactory memory of dogs.

### 2.2 Sniffer dogs’ characteristics

Dogs involved in the study were three belonging to the SecurityDogs a brand of NGS srl. Security company (Italy) one male and two females from three different breeds: Black German Shepherd, German Shepherd and Dutch Shepherd. The demographic characteristics of dogs are reported in Table 1.

**Table 1.** Demographic characteristics of dogs involved in the study protocol.

| Dog's name | Age (year) | Gender | Specie | Explosive substances trained |
| --- | --- | --- | --- | --- |
| HARLOCK | 3 | Male | Black German Shepherd | Yes |
| ROMA | 4 | Female | Dutch Shepherd | Yes |
| IDRA | 1 | Female | German Shepherd | No |

### 2.3 Materials used for sweat samples collection and training

Gauze was the elective support chosen for sweat collection, for its common distribution and consequent easy availability. The gauze used belong to Class IIa surgical device for its specific characteristics to be sterile, 100% cotton, latex and phtalate free (**Figure 1A**). Gauze with sweat were collected in glass jar with metal top (**Figure 1B**). The sniffer stand used for olfactory conditioning of dogs and the detection boxes used for the line-up are made of inhert materials to avoid plastics or adhesive materials that could be confoundent for the dogs’sniff. In each phase of dog training cross-contamination is avoided from sweat collection, storage, transport to training procedure. Training tools are projected to garantee that sweat samples never come in direct contact with dogs and tools are carefully sanitized at the end of each training session.

**FIGURE 1.**
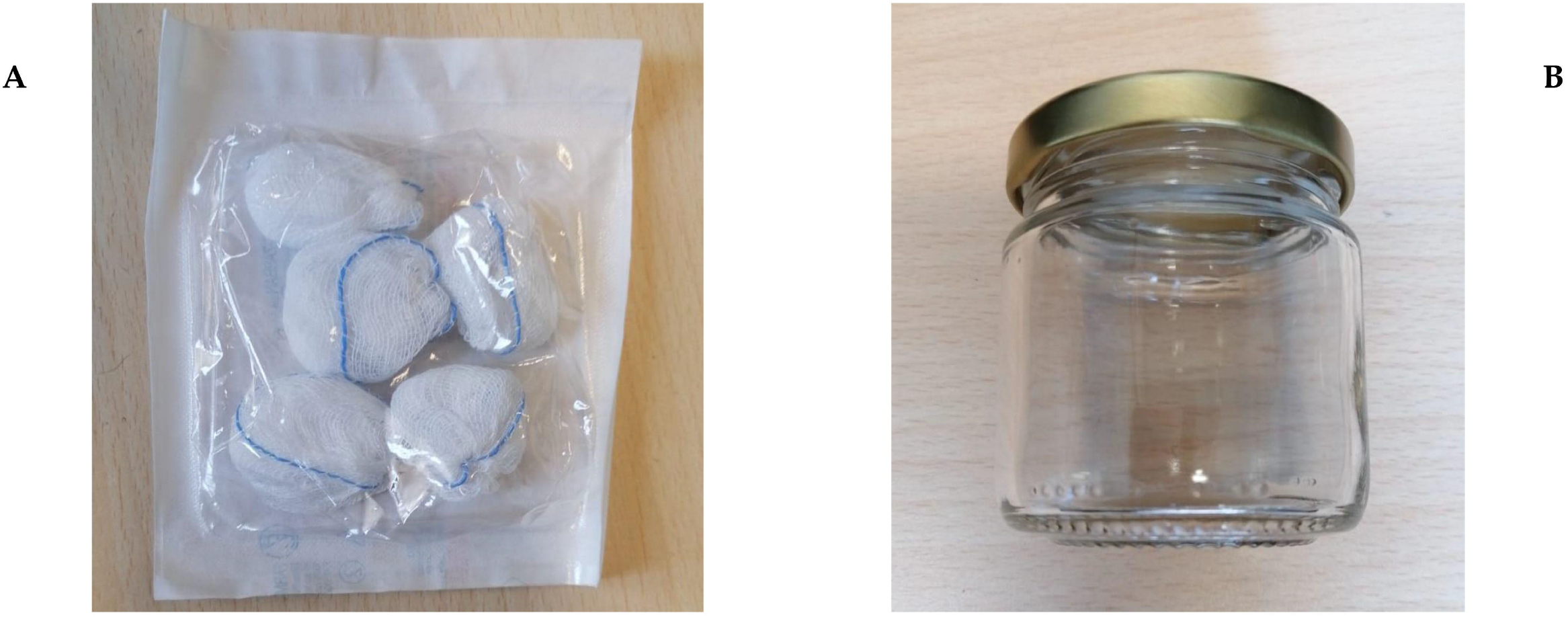
Gauze used for underarm sweat collection by patients (A). Glass jar with metal top used for gauze collection (B).

### 2.4 Sweat samples collection procedure

Skin sweat samples from patients with Covid-19 disease are collected at the Covid Center of the University Hospital Campus Bio-medico of Rome, while skin sweat samples from patients without Covid-19 disease collceted at the Internal Medicine Department of the University Hospital Campus Bio-medico of Rome. The demographic and clinical chracateristics of patients enrolled in the study are reported in Table 2.. Samples are anonimous identified and data registered on a database accessible only to the Principal Investigator of the study. Sweat samples are collected by patients with the assistance of the healthcare staff (physician or nusery) instructing them about the procedure, after informed consent has been recovered. This consists of self-collection by inserting gauze in the underarms, one for side, and mantaining in place for five minutes. After this, patients put both gauzes in the same glass can closed by the metal top and give it to the haelthcare staff that provides to the storage in a double bag for biological samples collection that can be delivered ouside the covid center to the Laboratory Division Glass can with sweat samples are delivered to the dog’s training space adhibited within the Drive-in campus test area of the University Hospital campus Bio-Medico of Rome within two hours from collection. If training is delayed samples are stored in refrigerate controlled tempertaure at 4°C for a maximum of 24 hours.

**Table 2.** Demographic and clinical chracateristics of patients enrolled in the study.

|  | Positive COVID-19 patients<br>n=20 | Negative COVID-19 patients<br>n=15 |
| --- | --- | --- |
| Mean Age (years) | 57.8 | 73.4 |
| Sex n (%) |  |  |
| Male | 9 (45) | 10 (67) |
| Female | 11 (55) | 5 (33) |
| Comorbidity n (%) | 9 (45) | 15 (100) |
| Diabetes | 2 (22) | 0 |
| High blood pressure | 6 (66) | 11 (73) |
| Cardiovascular disease | 0 | 11 (73) |
| Obesity | 3 (33) | 0 |
| Cancer | 3 (33) | 6 (40) |
| Ictus | 1 (11) | 0 |
| COPD | 0 | 4 (26) |
| Healthy n (%) | 11 (55) | 0 |
| Diagnosis at admission |  |  |
| COVID-19 pneumonia | 20 (100) | 0 |
| Pneumonia no Covid-19 | 0 | 6 (40) |
| Cardiovascular disease | 0 | 3 (20) |
| Ascites | 0 | 1 (6.6) |
| Diverticulitis | 0 | 1 (6.6) |
| Pleural effusion | 0 | 2 (13) |
| Sepsis | 0 | 2 (13) |

### 2.5 Dog training description

During training all operations of samples managment are performed by healthcare staff adequately equipped with personal protective equipment (PPE) including single-use water-repellent lab coats to avoid, FFP3 disposable mask, water-repellent cover shoes, nitrile gloves and face shield. The same PPE are also used by the dog trainers in any phase of the training. Dogs are equipped with working dog equipment including dedicated collars and gears daily sanitized.

Training is performed indoor in a dedicated and well equipped space conisiting in a container located within the Drive In Covid test area of the University Hopsital Campus Bio-medico of Rome. Local temperature is controlled and manitained within a temperature range of 20-25 °C to minimize influence on the experimental assay. The same temperature range is checked and requested also for sweat samples used for dog’s training.

Sweat samples collected in the glass jar after removal of the metal top are put inside the metal boxes used for line-up set-up and placed for 5 minutes before test start, time required for trace “ageing”. After this interval time, the sweat sample can be sniffed by dogs and the test begins. At the end of the test, lasting about 2 minutes and 30 seconds, boxes with the sweat sample are alternatively moved within the line-up and sanitized for dogs rotation.

During the specific conditioning to COVID-19 VOCs, an ad hoc designed sniffer stand has been used allowing the safe accomodation of the sweat sample and the comfortable positioning for the dog’s trainer when he enforces the the correct behaviour of the dog by the clicker and the material reward. In this phase is fundamental to use various sweat samples collected form different Covid-19 positive patients to fix in the olfactory memory of dog the presence of VOCs specific for Covid-19 within the vast range of odour emanations of the skin sweat. For the specific conditioning sniff repeats have been performed with positive reinforce by material rewards at each sniffing.

During the olfactory discrimination research the training aims at the detection of COVID-19 VOCs towards dog has been conditioned and the discrimination from what it is not of interest, that has to be extinguished. In this phase, detection boxes with sweat samples from COVID-19 positive patients, sweat samples from COVID 19 negative patients, blank gauze and empty box have been used. Sweat samples hide from the beginning and never visible at the dogs are positioned randomly by the help of a randomic software.

The basis of this training phase is in the handler-dog K-9 unit, where the handler realizes that the dog is ready for the autonomous detection through the line-up and for the correct and univocal response by “sitting” or “lying down”. This training phase is repeated from ten to tewenty times to fix the correct behaviour, depending by the dog’s ability. At this phase follows the verification of the training procedure by dedicated trial sessions for each dog during which

The number of correct and uncorrect identification of COVID-19 positive sweat samples was recorded for further statistical analysis of data.

### 2.6 Statistical analysis

The percentage difference between correct and uncorrect identification registered in the verification of the protocol procedure for each dog was evaluated by □^2^ test for proportions. p value <0.05 was considered statistically significant.

### 2.7 De-briefing session

Each training session has been video-recorded (as in the supplementary material) for further analysis during the de-briefing section performed at the end of each training session. Moreover, a specific report has been made for the training sessions tracing.

### 2.8 Sanitizing

Training site and training equipment were daily sanitized at the end of each training session, while waste disposal in special waste containers.

### 2.9 Troubleshooting

Sweat self-collection by inserting gauze in the underarms of the patients has to be carefully performed, as previously described to avoid any influence on training.

## 3. RESULTS

In April 2021 was completed the 4 weeks intensive training including 227 sessions and 700 tests with 92 different biological samples. These sessions aimed to identify the moment of switch for the dog. The switch is the time frame where the dog pass from a not relevant to a relevant percentage of correct reporting that has been fixed to 80% to be comparable to the gold standard diagnostic molecular and antigenic SARSCoV2 tests. From the switch, the training focused on fixing the correct behaviour otherwise the most reliable reporting by the dog, as much as possibile near to 100%. The gradual progression of the dogs in these sessions until the switch moment has been schematized in **Figure 2**. Exactly, after assigning a coefficient of difficulty for each training session that is directly proprotional to the number of boxes in the line-up, it was observed that the dog Harlock from a minumim of 46% of correct reporting reached a maximum of 92%, the dog Roma from a minimum of 63% arrived to 92% and the dog Idra from 53% pass to 100% of correct reporting. The occurred dog switch was evidenced in further trials sessions for each dog, exactly 17 for Harlock, 20 for Roma and 23 for Idra as reported in Table 3. Harlock correctly identied the COVID-19 positive sweat samples in the line-up 15/17 (88%), Roma 17/20 (85%) and Idra 20/23 (87%) times. The difference between the percentage of correct and uncorrect identifications was statistically significant (p<0.0001) (Table 1), confirming the switch obtained for each dog.

**Table 3.** Number (n°) and percentage (%) of total trials performed and correct and uncorrect identification by the dogs for the verification of the training protocol.

| Dog's name | Trial n. | Correct n° (%) | Uncorrect n° (%) | $\chi^2$ | p value |
| --- | --- | --- | --- | --- | --- |
| HARLOCK | 17 | 15 (88) | 2 (12) | 19.06 | <0.0001 |
| ROMA | 20 | 17 (85) | 3 (15) | 19.11 | <0.0001 |
| IDRA | 23 | 20 (87) | 3 (13) | 24.64 | <0.0001 |

**FIGURE 2.**
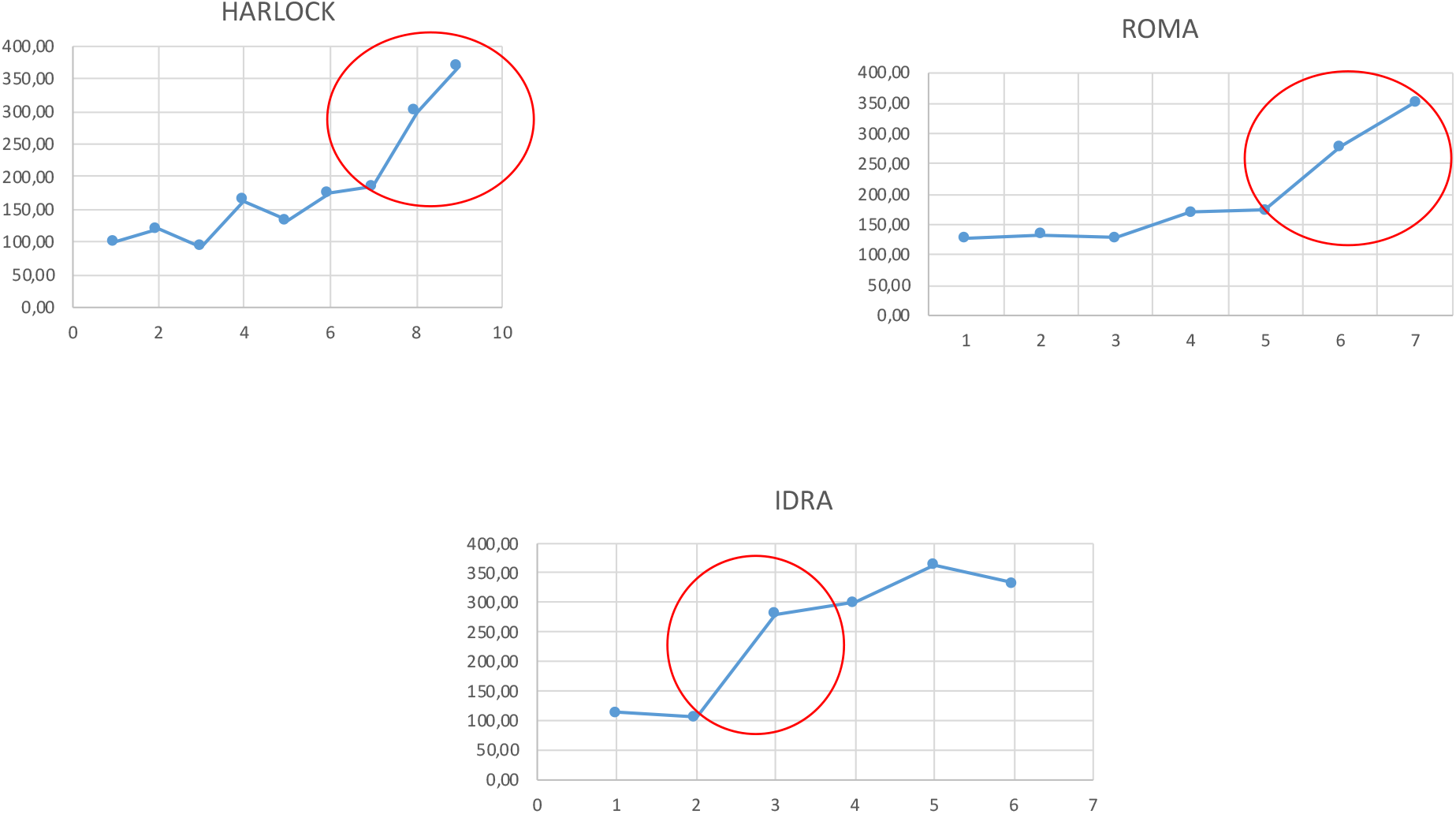
The gradual progression of the dogs until the switch moment (red cicle): percentage of correct identification normalized for the coefficient of difficulty (Yaxis) and number of trials (X axis).

## 4. DISCUSSION

Usually the test used for a mass screening mass should be rapid, enough sensitive, easy to manage, cheap and not time-consuming. The use of olfactory dogs has been proposed as fast, reliable and not expensive tool at this aim. The most critical factor is to provide a training protocol for sniffer dogs that could be easy to perform and enough reliable. The protocl prposed in this study provided some causes for reflection about the understanding of the “switch” moment for the dog. This moment corresponds to the time where the dog pass from a not relevant to a relevant percentage of correct reporting o be comparable to the gold standard diagnostic SARSCoV2 tests. In this study, data on the switch have been collected and the occurred dog switch evidenced in further trials sessions for each dog, with promising results in terms of sensitivity and specificty. Now the dog is ready for the mass screening in real-life. Overall, the proposed protocol, with focus on the switch moment, Could represent a valid support for sniffer dogs training replication in any setting.

The application of the proposed protocol for COVD-19 dog alert by sniffing axillary sweat samples, confirmed the ability of dogs, after specific training to detect COVID-19 VOCs. This approach provides a promising tool for COVID-19 mass screening at airports, stadium, or in case of crowded events where the virus transmission control by asymptyomatic individuals is fundamental in puclic health. After this first step, future perspectives will include training of further dogs using odor-less supports for skin emanation collection, the comparison between sniffer dogs ability and molecular RT-PCR gold test for Covid-19 diagnosis in different settings as the Drive In and the use of SARS-CoV2 proteins for dogs training to direct viral particles instead of VOCs from sweat samples.

## Data Availability

Not applicable

## Conflict of interest

The authors declare that there are no conflict of interests.

## Authors Contributions

Silvia Angeletti, Massimo Ciccozzi: study design, data analysis and study supervision; Francesco Travaglino, Silvia Spoto, Maria Chiara Pascarella, Giorgia Mansi and Raffaele Antonelli Incalzi: patient clinical diagnosis and sweat samples collection; Marina De Cesaris, Marta Fogolari: Covid-19 laboratory test; Marta Giovanetti: data analysis; Silvia Sartea: Logistic at the Drive in Area; Davide Plescia and Massimiliano Macera: study design and dog training. All Authors contributed to the manuscript preparation

## Notes

### Competing Interest Statement

The authors have declared no competing interest.

### Author Declarations

The study was approved by the Local Ethic Committee of University campus Bio-Medico of Rome (PAR 17.21 OSS).

